# Anticholinergic Co-Medication Burden Does Not Predict Cardiac Toxicity in Adolescent Diphenhydramine Exposure: A Pharmacovigilance Study of the FDA Adverse Event Monitoring System

**DOI:** 10.64898/2026.10.11.26365234

**Authors:** Zeyu Dong, Noura Fadel, Shrivathsan Krishnan, Preston Cai

## Abstract

**Objectives:** Social media trends such as the "Benadryl Challenge" have increased adolescent misuse of diphenhydramine (DPH), an over-the-counter antihistamine with anticholinergic activity. Whether co-ingested anticholinergic medications compound cardiac risk is untested. Using the Anticholinergic Cognitive Burden (ACB) scale, this study examined whether co-medication ACB burden is associated with outcome severity and cardiac toxicity in adolescent DPH cases reported to the FDA Adverse Event Monitoring System (AEMS).

**Methods:** This study extracted all adolescent reports (age 13–19) mentioning DPH in any drug role from AEMS, yielding 3,168 cases from 217,507 deduplicated records. Given non-normality (Shapiro–Wilk, all p < 0.001), non-parametric tests were used. A multivariable logistic regression predicted cardiac toxicity; logistic and random-forest models were compared by cross-validated AUC.

**Results:** Co-medication ACB burden correlated positively with case severity (ρ = 0.142, p < 0.001) and differed by age group (H = 19.52, p < 0.001). However, burden and cardiac risk diverged: cardiac cases carried a lower burden (mean 0.55 vs 1.27) and were younger, and burden did not independently predict cardiac toxicity (OR = 0.927, 95% CI [0.841, 1.022], p = 0.128). Younger age was the strongest predictor (OR = 0.865 per year, p < 0.001); cardiac events occurred in 10.2% of 13–15-year-olds versus 4.9% of older adolescents.

**Conclusion:** Anticholinergic co-medication burden and cardiac risk dissociate: burden tracks overall severity, but cardiac events concentrate in the youngest, least co-medicated adolescents, a profile consistent with deliberate single-agent overdose rather than polypharmacy. A low co-medication burden should not reassure clinicians triaging young adolescents.

## 1. Introduction

Adolescent misuse of over-the-counter (OTC) medications has risen sharply, driven in part by social media trends such as the "Benadryl Challenge," which encourages teenagers to ingest supratherapeutic doses of diphenhydramine (DPH) to induce hallucinations [1,2,3]. This trend coincided with an 87.2% increase in adolescent DPH ingestions reported to US Poison Centers between 2014 and 2023, alongside hospitalizations, seizures, and deaths, including the widely reported death of a 13-year-old in 2023 [4, 5]. In overdose, DPH behaves as a potent anticholinergic and sodium- and potassium-channel blocker, producing tachycardia, QT-interval prolongation, ventricular arrhythmia, delirium, seizures, and, rarely, death [6, 7, 8]. Many adolescents also use other drugs with anticholinergic activity: antidepressants, antipsychotics, and attention-deficit/hyperactivity disorder medications, and in older adults, such anticholinergic polypharmacy is associated with increased cardiac and neurological toxicity [9, 10].

These observations motivate an intuitive hypothesis: that adolescents carrying a greater cumulative anticholinergic burden from co-medications should be at higher risk of DPH-associated cardiac toxicity. Yet this hypothesis has never been tested in adolescents, and the data needed to test it are scarce. The Anticholinergic Cognitive Burden (ACB) scale, which assigns each medication an integer value of 1–3 according to its anticholinergic activity [11], is a tractable way to quantify cumulative burden, but it was developed and validated to predict cognitive and functional decline in older adults [12]; its relevance to acute cardiac risk in adolescents is unknown.

Using the FDA Adverse Event Monitoring System, we test whether higher co-medication ACB scores are associated with greater adverse-outcome severity and an increased probability of cardiac events in adolescent DPH cases, adjusting for patient age and the number of co-administered drugs. Because real-world adolescent presentations frequently involve DPH alongside other medications in varied clinical roles, this study deliberately captured the full spectrum of co-exposure rather than restricting attention to cases in which DPH was the sole suspected agent (Section 2.1). The cohort assembled here overturns the intuitive hypothesis: anticholinergic burden and cardiac risk dissociate, with implications for emergency triage and prevention.

## 2. Methods

The complete analysis pipeline, all intermediate files, and the statistical code are publicly available at https://github.com/AlexDongzeyu/Benadryl_Misuse.

### 2.1 Data source and cohort selection

This study analyzed adverse-event reports from the FDA Adverse Event Monitoring System (AEMS; formerly the FDA Adverse Event Reporting System, FAERS), a publicly available, de-identified pharmacovigilance database maintained by the U.S. Food and Drug Administration [13]. All available quarterly archives were downloaded and parsed into their demographic, drug, reaction, and outcome tables. To remove duplicate submissions of the same case, only the most recent report identifier per case identifier was retained.

The deduplicated demographic file was filtered to adolescent cases meeting three concurrent criteria: age 13–19 years inclusive, age reported in years, and country of occurrence recorded as the United States or unknown. This produced 217,507 unique adolescent records. DPH cases were then identified by matching 17 brand and generic names (e.g., diphenhydramine hydrochloride, Benadryl, Nytol, ZzzQuil, and Unisom) and by substring matching on "DIPHENHYDRAMIN," applied to both the reported drug name and the active-ingredient field to capture spelling variants and salt forms.

Cases were retained where DPH appeared in any reported drug role: primary suspect, secondary suspect, concomitant, or interacting. This inclusive definition was a design choice. Adolescent DPH presentations rarely occur in pharmacological isolation; restricting the cohort to reports in which DPH was the sole suspected agent would exclude the many real-world cases in which DPH is taken alongside other medications and would bias the cohort toward a narrow, artificially "clean" exposure that does not reflect clinical reality. Because the central question concerns the contribution of *co-administered* anticholinergic medications, a cohort that admits these co-exposures is both more representative and better matched to the hypothesis. The resulting analytic cohort comprised 3,168 cases (Fig. 1). Ages were grouped into early (13–15), middle (16–17), and late (18–19) adolescence to examine developmental differences in burden.

**Fig. 1.**
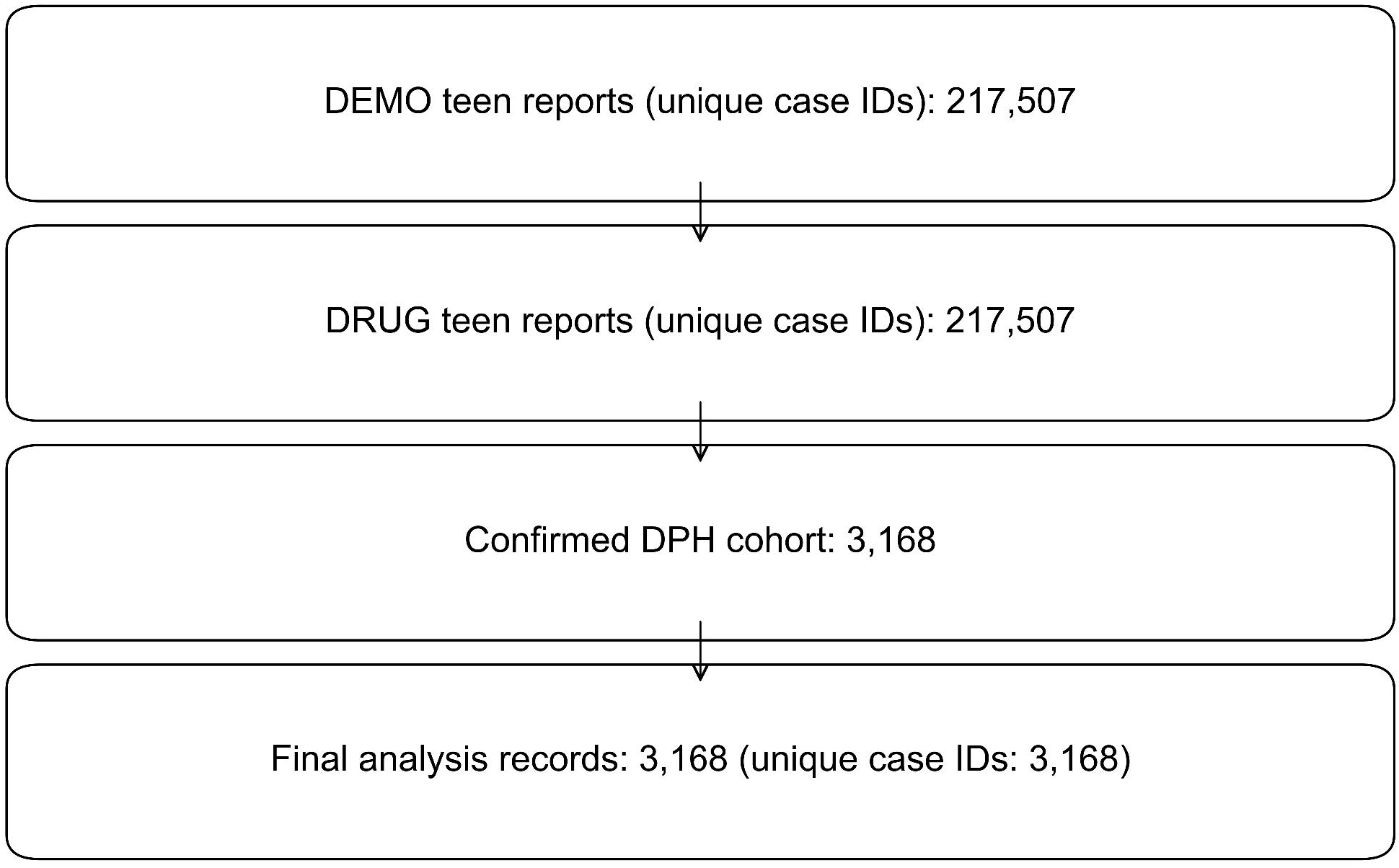
Cohort selection. From 217,507 deduplicated adolescent AEMS records, cases with confirmed DPH involvement in any reported drug role were retained, giving a final analytic cohort of 3,168.

### 2.2 Exposure and covariate construction

Co-administered drug names were standardized to ingredient level using the RxNorm RXNCONSO reference (ingredient terms only); unmapped drugs were retained under their reported names. Each drug was assigned an ACB score of 0–3 from a manually compiled table derived from published literature [11]. Two composite exposures were computed per case: total ACB including DPH (total_acb_with_dph) and total ACB from co-medications only (total_acb_codrugs_only). The co-medication score was the primary exposure because it isolates the anticholinergic contribution of concomitant medications from DPH itself (DPH alone scores 3). The number of co-administered drugs (n_codrugs) was retained as a covariate, and a binary indicator marked cases occurring after the September 2020 FDA safety communication [14].

### 2.3 Outcomes

Case severity was mapped from AEMS outcome codes to an ordinal scale: death = 4, life-threatening = 3, hospitalization = 2, and required intervention/disability/other = 1, taking the maximum across a case’s outcomes. The primary outcome, cardiac_any, was a binary flag for any QT-related event (electrocardiogram QT prolonged or abnormal, or long QT syndrome) or severe arrhythmia (torsade de pointes, ventricular tachycardia, ventricular fibrillation, or cardiac arrest).

### 2.4 Statistical analysis

Normality of the five primary variables was assessed by the Shapiro–Wilk test; all departed significantly from normality (all p < 0.001), so non-parametric methods were used throughout (distribution diagnostics in Supplementary Figs. S1–S10). A Kruskal–Wallis test (with η² effect size and Dunn’s Bonferroni-corrected post-hoc tests) compared co-medication ACB burden across age groups; Spearman’s correlation related burden to severity; and a Mann–Whitney U test (with rank-biserial effect size) compared burden between cardiac and non-cardiac cases.

A multivariable binary logistic regression modeled cardiac toxicity:

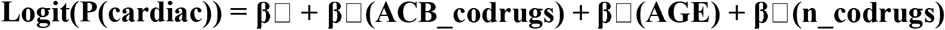

evaluated by McFadden’s pseudo-R², AIC, and AUC. With 219 cardiac events and three predictors (≈ 73 events per variable), the model comfortably exceeded the ≈ 10-events-per-variable guideline [15]. A formal age × ACB interaction term tested whether burden’s effect varied with age. To assess nonlinearity and obtain an unbiased performance estimate, logistic regression and a 500-tree random forest were compared under stratified five-fold cross-validation, with out-of-fold AUCs contrasted using DeLong’s paired test. Secondary analyses refitted the logistic model with age as a categorical group plus a pre/post-warning term and compared burden between sexes. Primary p-values were corrected for multiple comparisons by the Benjamini–Hochberg procedure. Analyses used Python (SciPy, statsmodels, scikit-learn, scikit-posthocs) and R (tidyverse, broom, pROC); α = 0.05.

## 3. Results

### 3.1 Cohort characteristics

The cohort comprised 3,168 adolescents with confirmed DPH involvement (Table 1). The mean age was 16.2 years (SD 1.9; median 16); 38.1% were aged 13–15, 32.0% aged 16–17, and 29.9% aged 18–19. The median co-medication ACB score was 0 (IQR 0–1; mean 1.22), the median total ACB including DPH was 3 (IQR 0–4), and cases involved a mean of 3.8 co-administered drugs (SD 5.3). Cardiac events were recorded in 219 cases (6.9%), and 22.6% of reports postdated the September 2020 FDA warning.

**Table 1.** Characteristics of the analytic cohort, overall and by cardiac outcome. Continuous variables are mean (SD) and median [IQR]; categorical variables are n (%). p-values compare cardiac vs non-cardiac cases (Mann–Whitney U for continuous, χ² for categorical).

| Characteristic | Overall (N = 3,168) | Non-cardiac (n = 2,949) | Cardiac (n = 219) | p-value |
| --- | --- | --- | --- | --- |
| Age, years | 16.2 (1.9); 16 [15–18] | 16.2 (1.9); 16 [15–18] | 15.7 (1.8); 15 [14–17] | < 0.001 |
| Age group, n (%) |  |  |  | < 0.001 |
| 13–15 | 1,208 (38.1) | 1,085 | 123 (10.2% cardiac) |  |
| 16–17 | 1,014 (32.0) | 964 | 50 (4.9% cardiac) |  |
| 18–19 | 946 (29.9) | 900 | 46 (4.9% cardiac) |  |
| Co-medication ACB burden | 1.2 (3.0); 0 [0–1] | 1.3 (3.1); 0 [0–1] | 0.6 (1.9); 0 [0–0] | < 0.001 |
| Total ACB incl. DPH | 2.9 (4.1); 3 [0–4] | 2.9 (4.2); 3 [0–4] | 2.5 (2.5); 3 [0–3] | 0.72 |
| No. of co-medications | 3.8 (5.3); 2 [0–5] | 3.9 (5.4); 2 [0–5] | 2.3 (4.3); 1 [0–2] | < 0.001 |
| Single-agent DPH (0 co-medications), n | 827 (26.1) | 744 (25.2) | 83 (37.9) | < 0.001 |
| (%) |  |  |  |  |
| **Maximum severity (0–4)** | 1.7 (1.5); 2 [0–3] | 1.6 (1.5); 2 [0–2] | 2.8 (1.0); 3 [2–4] | < 0.001 |
| **Post-warning era (≥ Sep 2020), n (%)** | 717 (22.6) | 712 | 5 (0.7% cardiac) | < 0.001 |

Compared with non-cardiac cases, cardiac cases were significantly younger, carried lower co-medication ACB burden, involved fewer co-medications, and were more often single-agent DPH reports (Table 1).

### 3.2 Anticholinergic burden increased with age

Co-medication ACB burden differed significantly across age groups (Kruskal–Wallis H = 19.52, p < 0.001, η² = 0.006; Fig. 3). Dunn’s post hoc tests (Bonferroni) showed that 13–15-year-olds carried a significantly lower co-medication burden than both 16–17-year-olds (p < 0.001) and 18–19-year-olds (p = 0.001); the two older groups did not differ (p > 0.99). Thus the youngest adolescents (those most associated with challenge-type misuse) entered the system with the least anticholinergic co-medication.

**Fig. 2.**
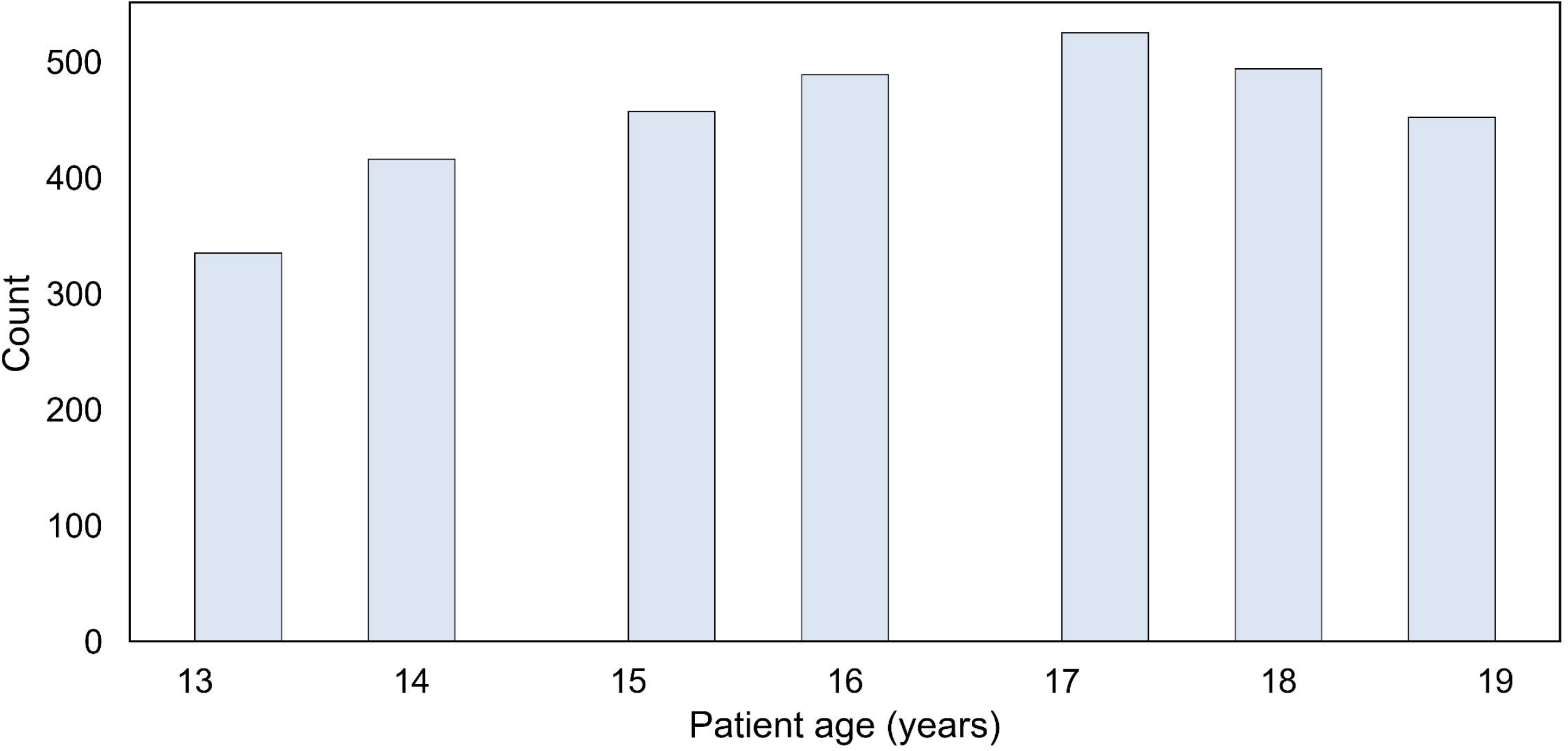
Age distribution of the analytic cohort (N = 3,168).

**Fig. 3.**
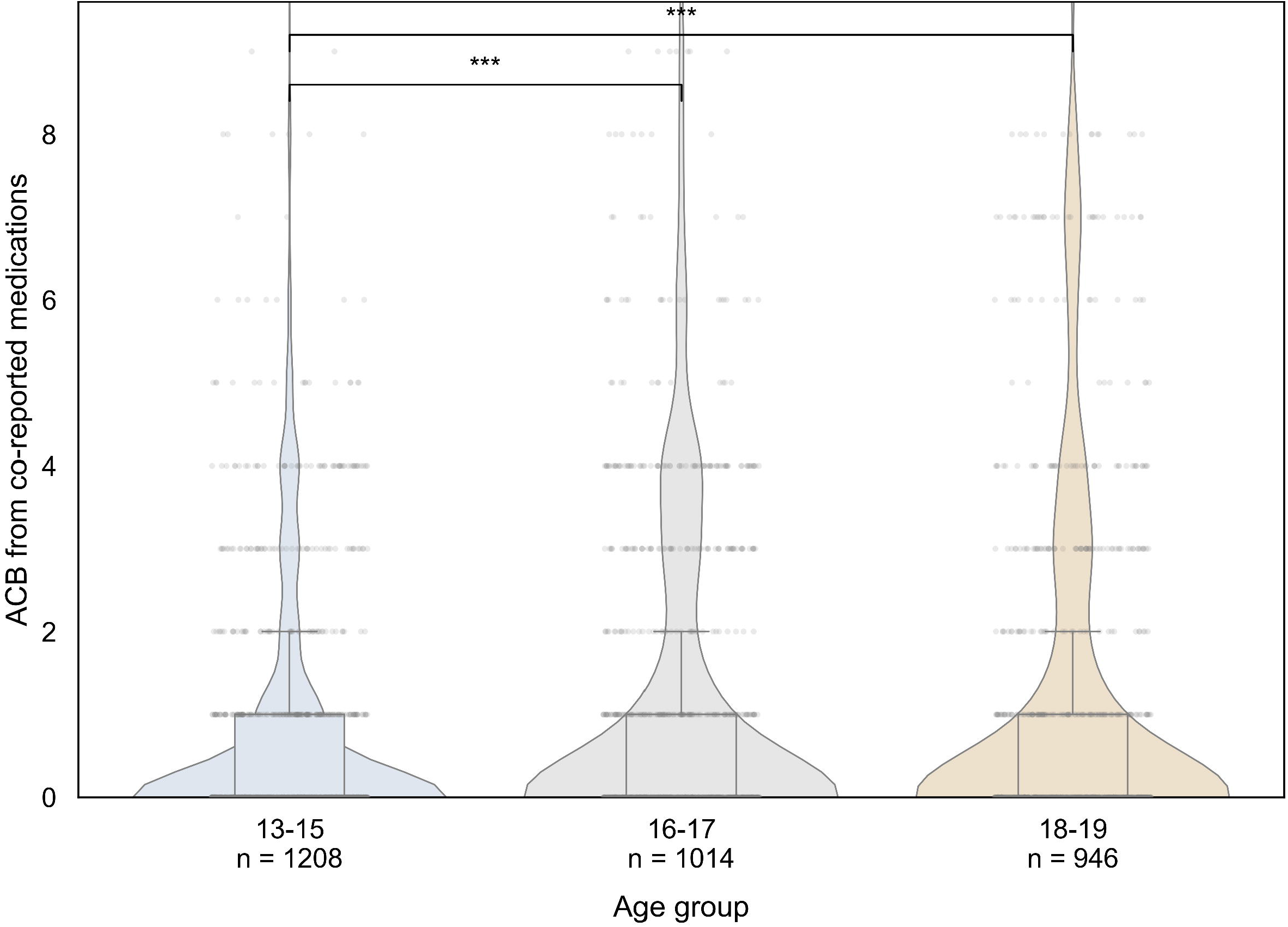
Co-medication ACB burden by age group (violin + box + jittered points). Kruskal–Wallis H = 19.52, p < 0.001, η² = 0.006. The 13–15 group carried significantly lower burden than the 16–17 (p < 0.001) and 18–19 (p = 0.001) groups; the older groups did not differ.

### 3.3 Burden-tracked overall severity

Co-medication ACB burden correlated positively and significantly with maximum case severity (Spearman ρ = 0.142, p < 0.001; Fig. 4). The smoothed relationship rose steeply between ACB scores of 0 and 3 and then plateaued, indicating that the severity gradient was steepest at low-to-moderate burden rather than strictly dose-linear.

**Fig. 4.**
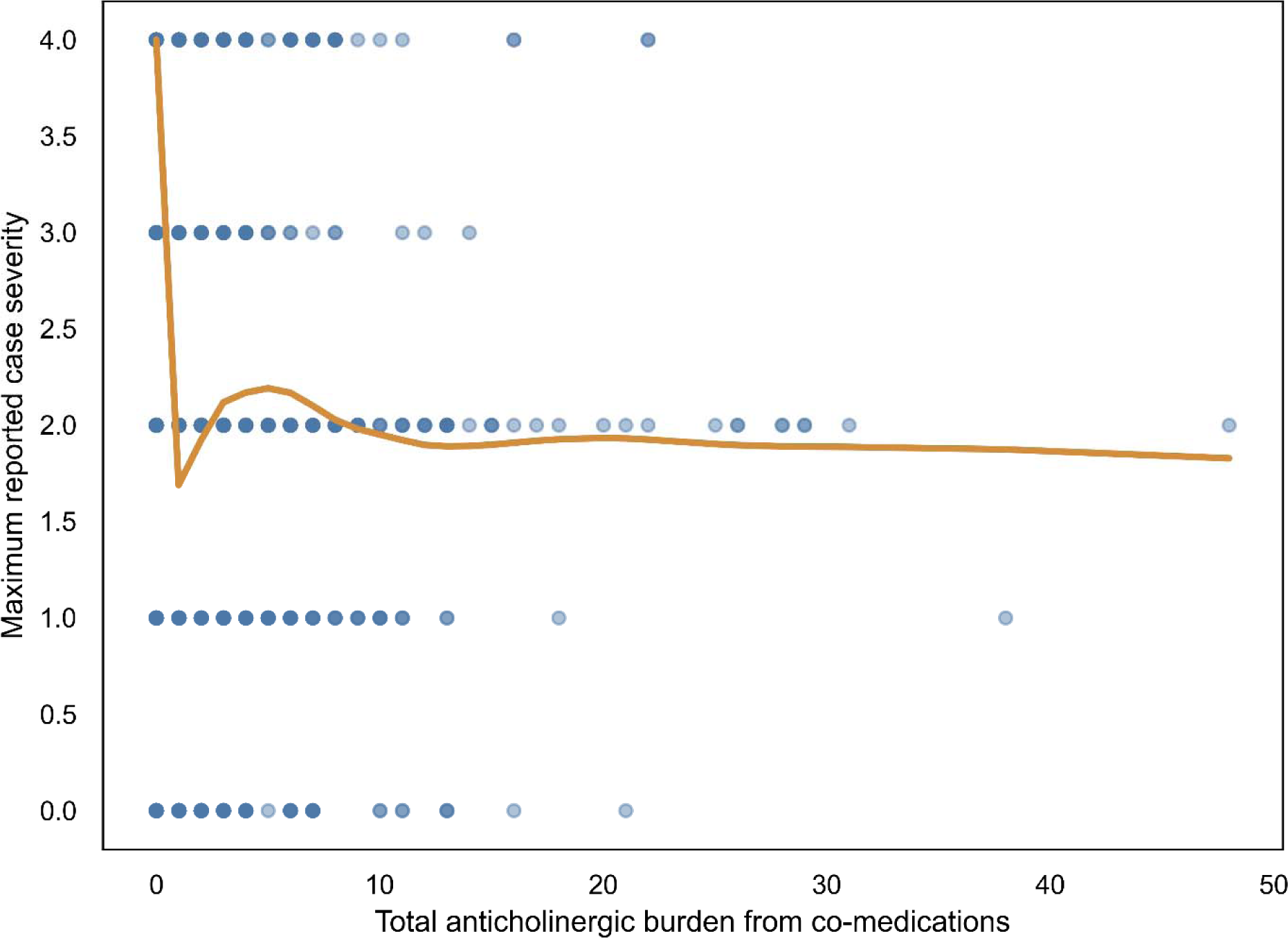
Co-medication ACB burden versus maximum case severity with a LOESS-smoothed trend (ρ = 0.142, p < 0.001).

### 3.4 Burden and cardiac risk diverged

In direct comparison, co-medication ACB burden differed significantly between cardiac and non-cardiac cases (Mann–Whitney U = 375,827, p < 0.001), but in the direction opposite to the severity finding: cardiac cases carried a lower burden than non-cardiac cases (mean 0.55 vs. 1.27; rank-biserial r ≈ −0.16). This inversion mirrored the cohort’s age structure. The youngest adolescents, who carried the least anticholinergic burden (Section 3.2), experienced the highest cardiac event rate (10.2% in 13–15-year-olds vs. 4.9% in both older groups; Table 1), and cardiac cases were more often single-agent DPH reports (37.9% vs. 25.2% with no co-medications) with fewer co-medications overall (median 1 vs. 2). Burden and cardiac risk therefore moved in opposite directions across the cohort.

### 3.5 Multivariable and machine-learning models

In the multivariable logistic model, co-medication ACB burden was not an independent predictor of cardiac toxicity (OR = 0.927, 95% CI [0.841, 1.022], p = 0.128; Table 2, Fig. 6). Younger age was the strongest predictor (OR = 0.865 per additional year, 95% CI [0.804, 0.930], p < 0.001), and the number of co-medications was also inversely associated with the cardiac outcome (OR = 0.921, 95% CI [0.877, 0.967], p = 0.001). The age × ACB interaction was non-significant (p = 0.17), indicating that burden’s (null) effect did not vary meaningfully with age and that the dominant signal was the age main effect. The model showed modest discrimination (apparent AUC = 0.652; McFadden’s pseudo-R² = 0.029; AIC = 1,553.9).

**Fig. 5.**
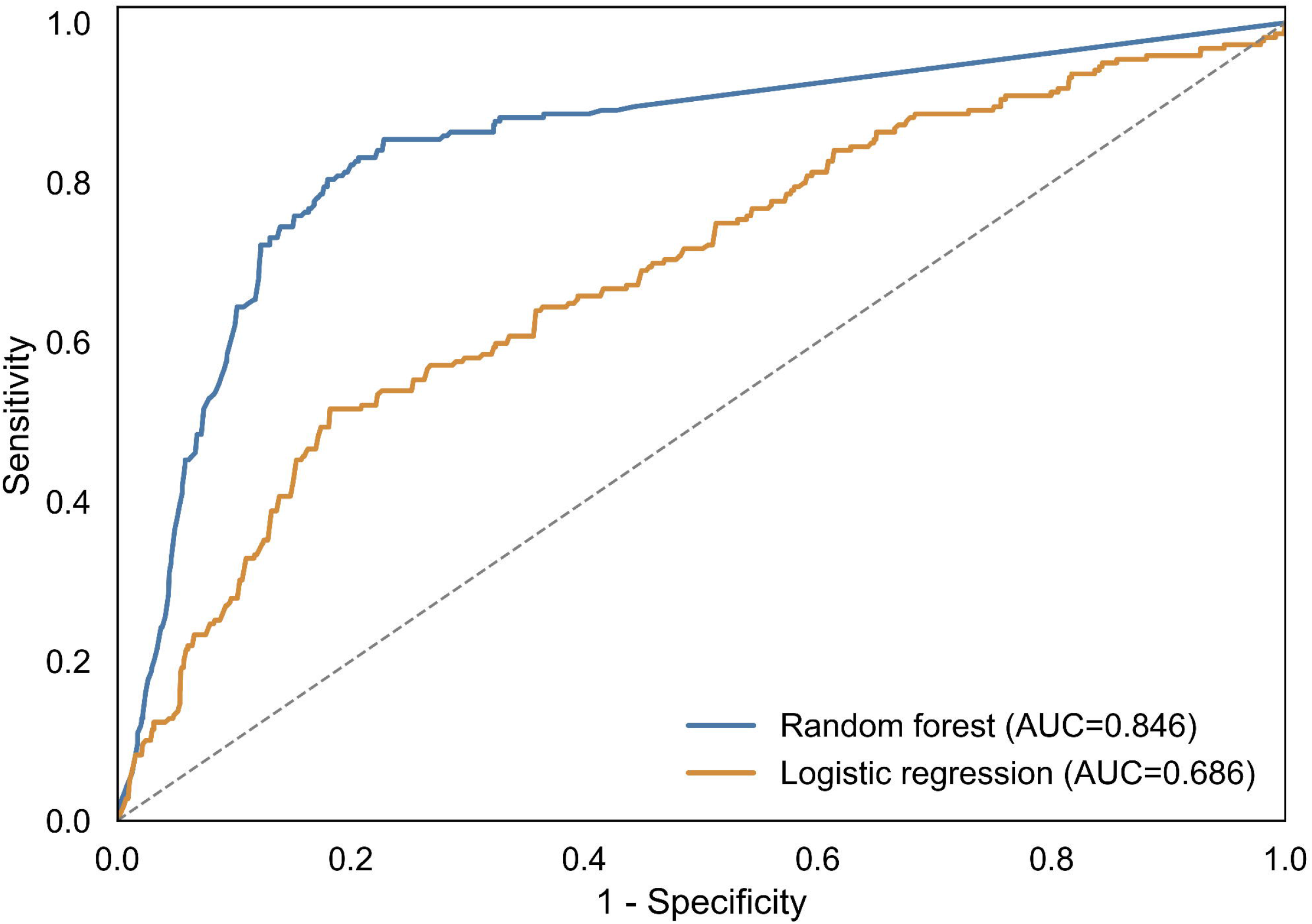
Cross-validated ROC curves. The random forest (AUC 0.769) significantly outperformed logistic regression (AUC 0.640; DeLong p < 0.001).

**Fig. 6.**
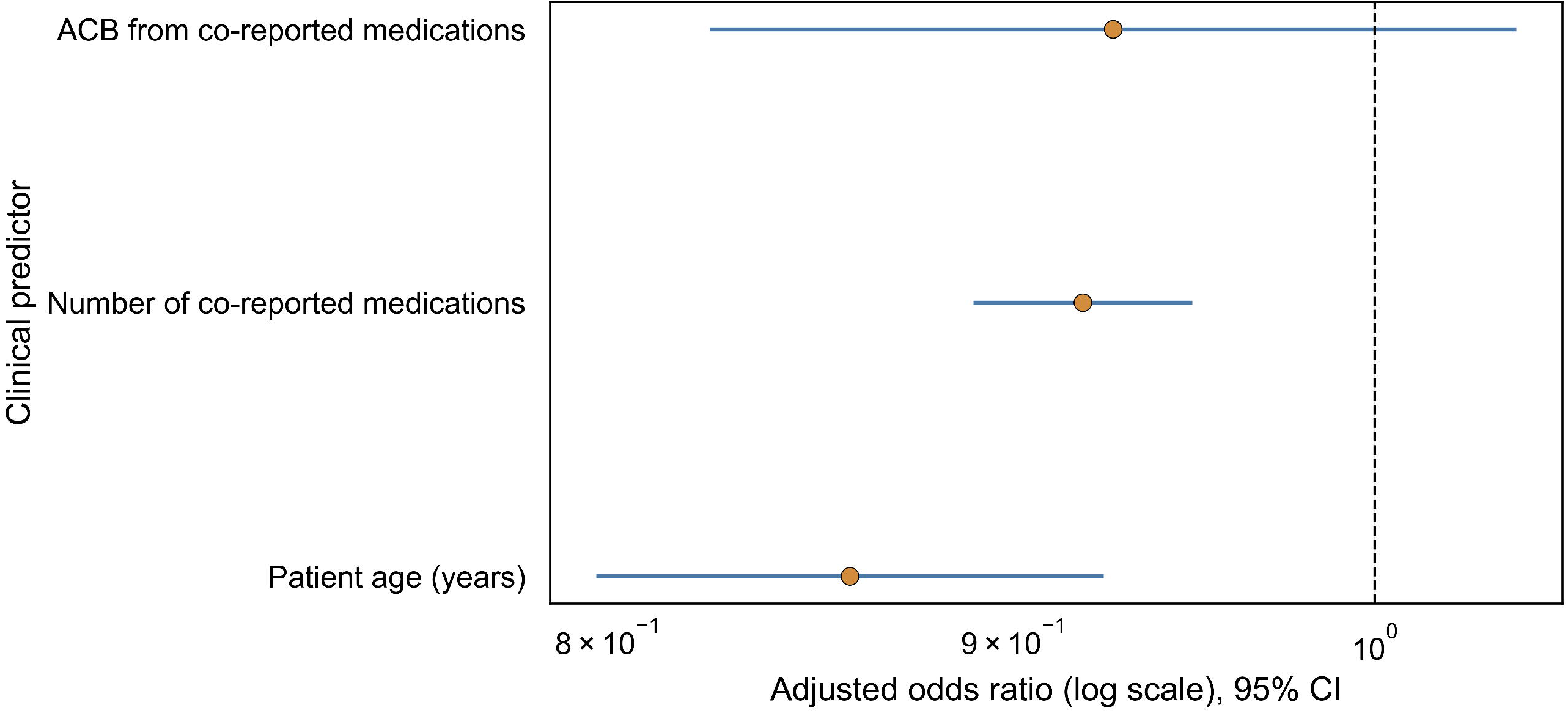
Adjusted odds ratios for cardiac toxicity. Age (OR 0.865, p < 0.001) and number of co-medications (OR 0.921, p = 0.001) were significant; co-medication ACB burden was not (OR 0.927, p = 0.128). Dashed line, OR = 1.

**Table 2.** Multivariable logistic regression for cardiac toxicity (N = 3,168; 219 events). Model: cardiac_any ∼ co-medication ACB + age + number of co-medications.

| Predictor | Adjusted OR | 95% CI | p-value |
| --- | --- | --- | --- |
| Co-medication ACB burden (per point) | 0.927 | 0.841–1.022 | 0.128 |
| Age (per year) | 0.865 | 0.804–0.930 | < 0.001 |
| No. of co-medications (per drug) | 0.921 | 0.877–0.967 | 0.001 |

Under cross-validation, a random forest substantially outperformed logistic regression (AUC 0.769 vs. 0.640; DeLong p < 0.001; Fig. 5), indicating that nonlinear, interaction-laden structure among the predictors carries discriminative information the additive logit cannot capture. Because both models were re-weighted toward the rare positive class, their probability estimates were not well calibrated (Supplementary Fig. S11); the random forest is therefore reported as a discrimination benchmark rather than a deployable calculator.

### 3.6 Secondary analyses

Co-medication ACB burden did not differ by sex (Mann–Whitney p = 0.176, rank-biserial r = −0.025; n = 2,760; Supplementary Fig. S12). The age-categorical sensitivity model reproduced the protective age gradient (16–17 vs 13–15: OR 0.473; 18–19 vs 13–15: OR 0.450; both p < 0.001). A temporal term showed markedly fewer coded cardiac events after the 2020 warning (0.7% vs. 8.7%); given the magnitude and the instability of the post-warning subgroup, this study treats this as an exploratory, likely reporting-related signal rather than a true decline in cardiotoxicity. All three primary tests and the DeLong comparison remained significant after Benjamini–Hochberg correction; the sex comparison did not.

## 4. Discussion

Across 3,168 adolescent AEMS reports, the largest such cohort to date, anticholinergic co-medication burden and cardiac toxicity risk moved in opposite directions. Burden was positively associated with overall reported severity, yet cardiac events concentrated in the youngest adolescents, precisely those carrying the least anticholinergic co-medication. This dissociation overturns the intuitive expectation that greater anticholinergic polypharmacy means greater cardiac danger, and it changes how adolescent DPH presentations should be triaged.

### 4.1 Two distinct exposure phenotypes

Two distinct exposure phenotypes within the cohort best explain the findings. The first is a deliberate single-agent overdose phenotype: younger adolescents ingesting large doses of DPH alone, consistent with challenge-type or self-harm misuse, characterized in our data by low co-medication burden, few co-medications (37.9% of cardiac cases involved DPH alone), and the highest cardiac event rate. The second is an anticholinergic polypharmacy phenotype: older adolescents on multiple, often psychiatric, medications [9, 16], characterized by higher ACB burden but lower cardiac event rates, which probably reflect supervised therapeutic exposure rather than massive deliberate ingestion.

This interpretation matches the largest prior poison center analysis of adolescent DPH ingestions, which found that intentional ingestions, and suspected suicide in particular, carried substantially higher rates of cardiac complications, seizures, and death than misuse or abuse [17]. Because DPH cardiotoxicity is dose-driven through hERG-channel blockade [7] and is otherwise uncommon owing to the drug’s wide therapeutic range [18], acute cardiac risk most likely reflects dose-escalating single-agent overdose rather than the number or anticholinergic burden of co-medications.The positive burden–severity association (Section 3.3) is consistent with anticholinergic co-medications compounding overall toxicity, but this does not extend to the specific, dose-dependent cardiac endpoint.

### 4.2 The age paradox

Younger age was the dominant cardiac risk factor after adjustment (OR = 0.865 per year, p < 0.001), with 13–15-year-olds experiencing roughly double the cardiac event rate of older adolescents despite carrying the lowest anticholinergic burden. Two mechanisms plausibly converge. Biologically, peripubertal differences in cardiac electrophysiology may heighten susceptibility to drug-induced QT prolongation in early adolescence [19,20,21], increasing sensitivity to DPH’s hERG-channel blockade [7]. Behaviorally, younger adolescents are more prone to challenge-based, risk-taking misuse owing to ongoing cognitive development [22]; the reported death of a 14-year-old with a postmortem DPH concentration of 49,658 ng/mL following the challenge illustrates the dose escalation possible in this group [23]. That the age × ACB interaction was non-significant (Section 3.5) argues that age is an independent driver rather than a modifier of any anticholinergic effect.

### 4.3 Modeling and signal structure

The superior cross-validated performance of the random forest over logistic regression (AUC 0.769 vs. 0.640; DeLong p < 0.001) indicates that the relationship between these predictors and cardiac toxicity is nonlinear and non-additive, as expected if a dose-escalating, age-dependent phenotype, rather than a smooth additive burden gradient, underlies the risk. The deliberate re-weighting toward the rare positive class improved discrimination at the cost of calibration, so these models are best read as evidence of signal structure and as hypothesis-generating tools, not as clinical risk calculators.

### 4.4 Clinical implications

In adolescent DPH presentations, a low anticholinergic co-medication burden should not be taken as reassurance, particularly in younger teenagers. The cardiac-risk signature in this cohort was youth combined with single-agent, presumably high-dose, DPH ingestion. Prevention and triage messaging that focus on polypharmacy or that assume "more drugs, more danger" may therefore miss the highest-risk group. Consistent with prior calls to prioritize the suicide-attempt pathway over misuse alone [17], clinicians and poison centers should focus age-targeted vigilance on deliberate single-agent overdose.

## 5. Limitations

Several limitations apply. First, AEMS relies on voluntary reporting and preferentially captures severe cases; co-ingested drugs are likely under-reported, so the true anticholinergic burden may be underestimated, and the database supports association rather than causation. Second, AEMS does not record intent, dose, or serum concentration, so the single-agent overdose phenotype is inferred from co-medication structure rather than measured directly; linkage to poison center or clinical data would allow this inference to be tested. Third, co-medication burden and the number of co-medications are strongly collinear (Spearman ρ ≈ 0.61), so their adjusted coefficients should be interpreted jointly. Fourth, the ACB scale was validated in older adults and may not capture anticholinergic risk in adolescents; a pediatric-specific scale would strengthen future work. Finally, the post-warning temporal subgroup was small and unstable, and the apparent post-2020 decline in coded cardiac events most likely reflects changes in reporting or coding rather than a true effect.

## 6. Conclusion

Across 3,168 adolescent AEMS reports, anticholinergic co-medication burden was associated with greater overall case severity but did not independently predict cardiac toxicity; instead, cardiac events concentrated in the youngest, least co-medicated adolescents. Burden and cardiac risk therefore dissociate, pointing to deliberate single-agent overdose, rather than anticholinergic polypharmacy, as the more likely driver of acute cardiac risk.Age-targeted prevention and prospective, clinically adjudicated studies are the next priority.

## Declaration of generative AI and AI-assisted technologies in the manuscript preparation and data analysis process

During the preparation of this work the authors used GitHub Copilot and Claude Code in order to improve the code, language and readability of the research. After using this tool, the authors reviewed and edited the content as needed and take full responsibility for the content of the publication.

## 8. Declarations

### Data availability

AEMS source data are publicly available from the U.S. FDA. All processed analysis tables and code are publicly available at https://github.com/AlexDongzeyu/Benadryl_Misuse

### Ethics approval

This study used publicly available, fully de-identified data from the FDA Adverse Event Monitoring System and did not involve any interaction with human participants or access to identifiable private information. It therefore did not require institutional review board approval or informed consent under regulations governing secondary use of de-identified public data.

## Acknowledgements

The authors thank Youreka Canada for its support and mentorship.

## Abbreviations

ACB: Anticholinergic Cognitive Burden
AEMS: Adverse Event Monitoring System
AIC: Akaike Information Criterion
AUC: Area Under the Curve
CI: Confidence Interval
DPH: Diphenhydramine
FAERS: FDA Adverse Event Reporting System
IQR: Interquartile Range
OR: Odds Ratio
OTC: Over-the-Counter
SD: Standard Deviation.

## 9. Supplementary material Supplementary figures

- **S1–S10. Distribution diagnostics** (histograms, Q–Q plots) for age, total ACB with DPH, co-medication ACB, number of co-medications, and severity: age_histogram.png, age_qq_plot.png, acb_with_dph_histogram.png, acb_with_dph_qq_plot.png, acb_codrugs_only_histogram.png, acb_codrugs_only_qq_plot.png, codrug_count_histogram.png, codrug_count_qq_plot.png, severity_histogram.png, severity_qq_plot.png.
- **S11. Calibration curves** (logistic vs. random forest, out-of-fold): calibration_curve_cv.png.
- **S12. Sex-stratified co-medication ACB burden:** sex_stratified_acb_violin.png.
- **S13. Drug co-occurrence network** (DPH as central hub): drug_cooccurrence_network.png.
- **S14. Random-forest SHAP feature importance:** shap_beeswarm.png.
- **S15. Co-medication ACB burden by cardiac outcome:** acb_by_cardiac_outcome.png.

